# Effect of Hypokalemia on Screening for Primary Aldosteronism

**DOI:** 10.1101/2022.06.16.22276370

**Authors:** Nicholas W Rizer, Xiruo Ding, Debbie L Cohen, Daniel S Herman

**Affiliations:** Department of Pathology and Laboratory Medicine, The Perelman School of Medicine, University of Pennsylvania; Department of Emergency Medicine, The Johns Hopkins School of Medicine; Department of Biomedical Informatics and Medical Education, University of Washington; Division of Renal, Electrolyte, and Hypertension, Department of Medicine, The Perelman School of Medicine, University of Pennsylvania

**Keywords:** Primary Aldosteronism, Hypertension, Secondary Hypertension, Screening, Hypokalemia

## Abstract

**Context:** Primary aldosteronism (PA) is a common, underdiagnosed, and treatable cause of hypertension. Guidelines recommend testing for hypokalemia, but current practices and their impact are unknown.

**Objective:** To estimate the impact of hypokalemia on the results of PA screening in clinical practice.

**Methods:** We studied electronic health records for adults who received longitudinal care within a large, integrated health system between 2007 and 2017 and underwent paired blood aldosterone and renin testing. Patients were grouped based on concurrent blood potassium concentrations.

**Results:** We found that 2,940 (82%) of the 3,571 patients screened for PA had concurrent potassium testing. Among all patients with a negative PA screen, 18% of patients did not have concurrent potassium testing and 14% of those tested were hypokalemic. Of the screen-negative hypokalemic patients, 77% were not retested. Among patients with repeated screens, we found that initial potassium concentration was a predictor of PA screen positivity on repeat. A one standard deviation lower initial potassium concentration was associated with a 74% higher odds of screen positivity on repeat, independent of initial aldosterone and renin results.

**Conclusions:** In-practice clinical data demonstrate that unrecognized and uncorrected hypokalemia contributes to the underdiagnosis of PA. Universal potassium testing and rescreening for PA in the setting of hypokalemia would likely substantially improve the identification and thereby management of PA.

## Introduction

Primary aldosteronism (PA) is the most common cause of secondary hypertension in the United States and is present in approximately 20% of patients with resistant hypertension. PA is associated with increased risk of left ventricular hypertrophy, myocardial infarction, atrial fibrillation and cerebrovascular events [1–6]. To reduce the conferred excess risk of morbidity and mortality, patients suspected to have PA should be screened for PA by measurement of serum or plasma aldosterone concentration, renin concentration or activity, and calculation of the aldosterone-to-renin ratio (ARR) [7]. Due to underdiagnosis of PA, multiple specialty societies’ guidelines recommend that all patients with resistant hypertension should be screened for PA and all patients with suspected PA should be referred to a hypertension specialist [7,8]. The limited available data indicates that these guidelines are poorly, or at least variably, integrated into clinical practices yielding lower than expected frequencies of PA screening and diagnosis [9–11].

The diagnosis of PA is challenging, in part, because of the complexities of administering and interpreting PA screening tests. Several factors have been shown to affect patient aldosterone and renin concentrations [12–15]. In an effort to control these potentially confounding factors, guidelines have suggested specific best practices for ensuring technically adequate and consistent testing conditions. Baseline blood potassium concentrations are important as hypokalemia, while a cardinal feature of the classic presentation of PA, can suppress aldosterone concentration and lead to false-negative PA screening results [16]. Classic physiology and more recent studies have shown that in healthy adults secretion of aldosterone is suppressed by low potassium and augmented by elevated potassium [17–19]. Blood potassium concentration is regulated by negative feedback via aldosterone, whereby blood potassium concentration increases adrenal cortical secretion of aldosterone which acts on the cortical collecting tubule of the nephron to increase secretion of potassium. Additionally, several other mechanisms exist for potassium regulation apart from aldosterone [20–22].

Guidelines recommend measurement of blood potassium for all patients being screened for PA and potassium supplementation for hypokalemic patients prior to PA screening [7]. However, the evidence base for this recommendation is limited and largely reliant on studies of few patients with severe PA who are not likely representative of the clinically screened population [23]. There is no in-practice literature supporting the recommendation for potassium supplementation prior to PA screening in patients with hypokalemia. In particular, there is no real-world evidence demonstrating a clinically important effect of hypokalemia on PA screening interpretation. This is important to consider, as universal potassium evaluation and repletion is a logistical obstacle to widespread PA screening. On the other hand, if potassium strongly affects aldosterone concentrations, without universal potassium testing and repletion many PA patients would be missed. To our knowledge, no study has comprehensively examined the frequency and utility of potassium testing at the time of PA screening. Thus, to inform future guidelines and approaches for detection and diagnosis of PA, retrospective electronic health record (EHR) data were used to characterize current screening practice within a large, diverse health system and examine the impact of hypokalemia on PA screening results.

## Methods

### Patients

The University of Pennsylvania Health System (UPHS) is a large academic health system with primary care and specialist outpatient sites and referral centers in southeastern Pennsylvania and southern New Jersey. Patient records were obtained from patients (N=488,412) within our health system who had received longitudinal care, defined as at least 5 office visits in at least 3 distinct years, between 2007 and 2017. We proceeded to examine 3,572 patients who had concurrent serum or plasma aldosterone and plasma renin activity testing. One patient was excluded because of missing documentation of sex.

For the purposes of this study the time of PA screening was used as the index date. For comparison purposes, we randomly selected a 10:1 control population of patients without aldosterone or renin testing, matched on sex and age-deciles at the time of a randomly chosen blood potassium measurement.

Several analyses were performed on two cohorts of patients: patients with initially negative PA screening tests (See Figure 1A) and patients with repeated PA screening and potassium testing grouped by blood potassium status trajectory (See Figure 1B). When examining patients with more than two PA screening tests, we restricted analyses to patients’ initial PA screening test and the immediately subsequent PA test. For analyses with respect to blood potassium concentrations (Figure 1B), we assigned trajectories based on potassium measurement at the time of initial and repeated PA screening tests. We excluded patients with hyperkalemia (N=212), a glomerular filtration rate (GFR) less than 45 mL/min prior to their initial PA screen (N=95), or interval adjustments in potassium sparing diuretics or mineralocorticoid receptor antagonists (N=2). All analyses were performed using custom python scripts available at https://bitbucket.org/hermanlab/pa_screening_hypokalemia. This study was reviewed and approved by the University of Pennsylvania Institutional Review Board (#827260).

**Figure 1.**
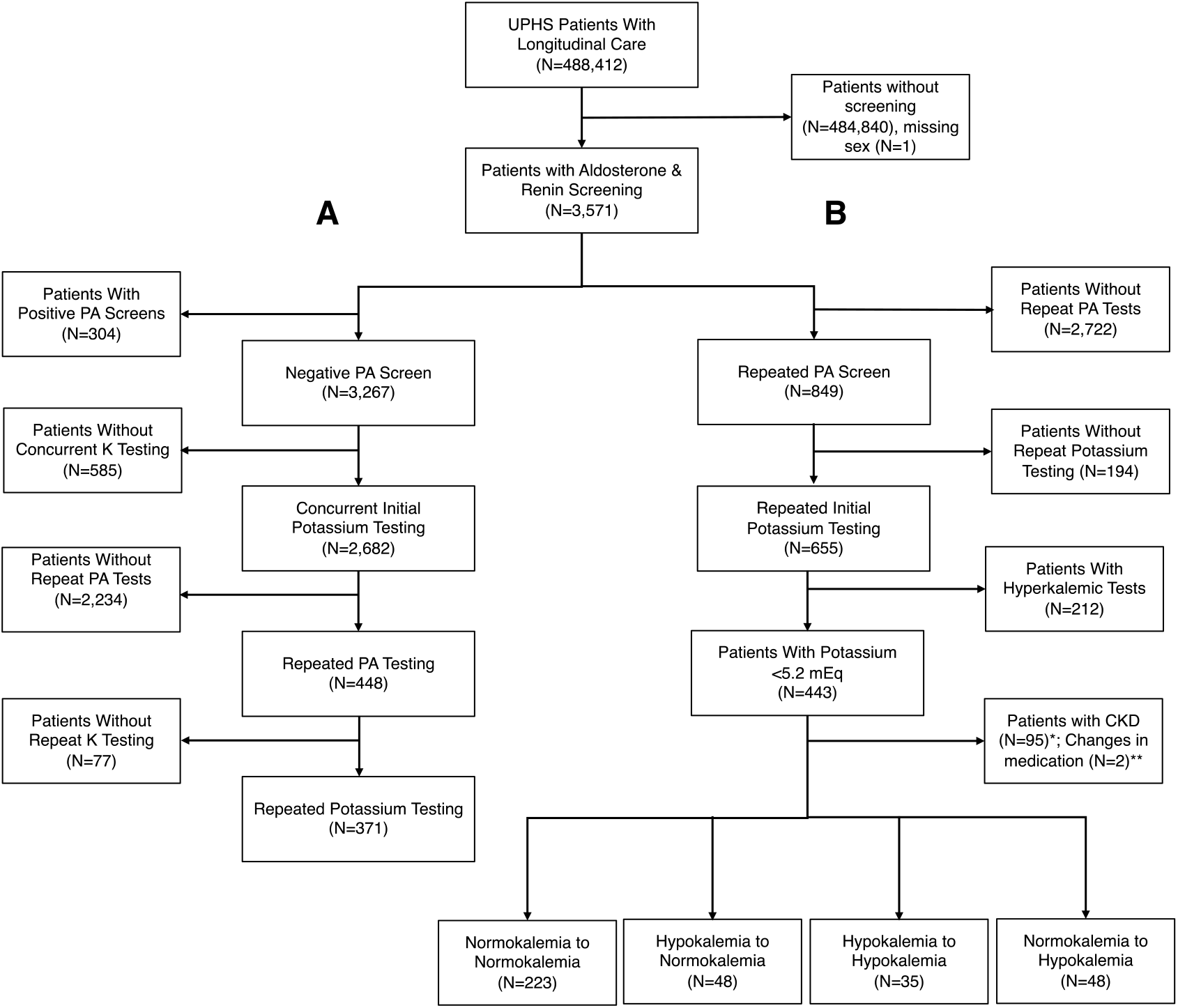
Study design. Flow diagram of cohort selection. (A) Cohort of patients used in analyses of repeated initially negative PA screening tests. (B) Cohorts of patients used in potassium trajectory analyses. *Patients with CKD Stage 3B or greater any time prior to first PA screening test. **Patients with addition or removal of mineralocorticoid receptor antagonist or potassium sparing diuretic.

### Clinical Data

EHR data were extracted from the Penn Data Store clinical data warehouse (on 4/14/2018) using custom python and R scripts (http://www.bitbucket.com/hermanlab/EHR_transform) and partially de-identified into a limited dataset. The extracted data included encounter meta information (visit type, blood pressure, location), laboratory testing results, medication prescriptions, diagnosis codes, and demographics. Intermediate data files were encrypted and de-identified. Data were left censored at January 1, 1997, filtered, cleaned, harmonized, and deposited in an SQLite (version 3.23.1) database. Subsequent analyses were restricted to outpatient encounter data.

### Classification of PA, Potassium, and Medication Status

To assess if a patient had a positive screen for PA, we applied conservative interpretive criteria used in our clinical practice that are expected to be highly clinically specific for PA: aldosterone ≥ 15 ng/dL, plasma renin activity < 0.5 ng/mL/hr, and ARR ≥ 30 [7,24,25]. Recent work has shown that an aldosterone concentration of approximately 15 ng/dL with suppressed renin was highly predictive of PA, further supporting these interpretive criteria [26]. To calculate the ARR, all renin activity values reported below the lower limit of the reportable range were conservatively set to the lower limit of the reportable range (e.g. < 0.1 ng/mL/hr set as 0.1 ng/mL/hr). We identified concurrent blood chemistry results as those from a specimen collected closest to the index or repeat screening date and no more than 30 days apart. Potassium results were classified as low (≤ 3.5 mEq/L), normal (3.6 mEq/L - 5.1 mEq/L), or high (≥ 5.2 mEq/L) based on the test’s reference interval. Patients were considered to be taking a selected medication if the prescription was initiated on or before the PA screening encounter and if the prescription was stopped on or after the same screening encounter.

### Statistical Analysis

The relationship between initial potassium concentration and repeat PA screening or PA screen positivity was assessed using univariate and multivariate logistic regression. Pre-specified covariates included the initial potassium concentration, the logarithm of the initial aldosterone concentration, the logarithm of the initial renin concentration, sex, race, and the time interval between PA screening tests. All continuous variables were standardized, and all covariate associations were reported as odds ratios (OR).

We assessed the relationship between within-patient changes in aldosterone and potassium concentrations using orthogonal linear regression over standardized delta values. The linear relationship was reported as unstandardized values. Categorical data were compared across classes using Fisher’s Exact tests. The changes in aldosterone concentrations across repeat examinations were compared across potassium trajectories using Mann-Whitney U-tests.

## Results

### Primary Aldosteronism Screening

To characterize PA screening practices in our health system, we retrospectively examined data from 488,412 patients who received longitudinal care in UPHS between 2007 and 2017. We identified 4,421 paired aldosterone and renin test results for 3,571 (0.7%) distinct patients (Figure 1 and Table 1). As compared to control patients never screened for PA and matched on age-deciles and sex, these patients were more likely to have a documented race of Black (46% versus 24%; p < 1.0×10^−10^) and had slightly lower blood potassium concentrations (4.1 mEq/L versus 4.3 mEq/L; p = 3.5×10^−4^). Of all screened patients, 354 (9.9%) patients met conservative PA criteria, including 304 patients (8.5%) at the time of their initial PA screen.

**Table 1.**
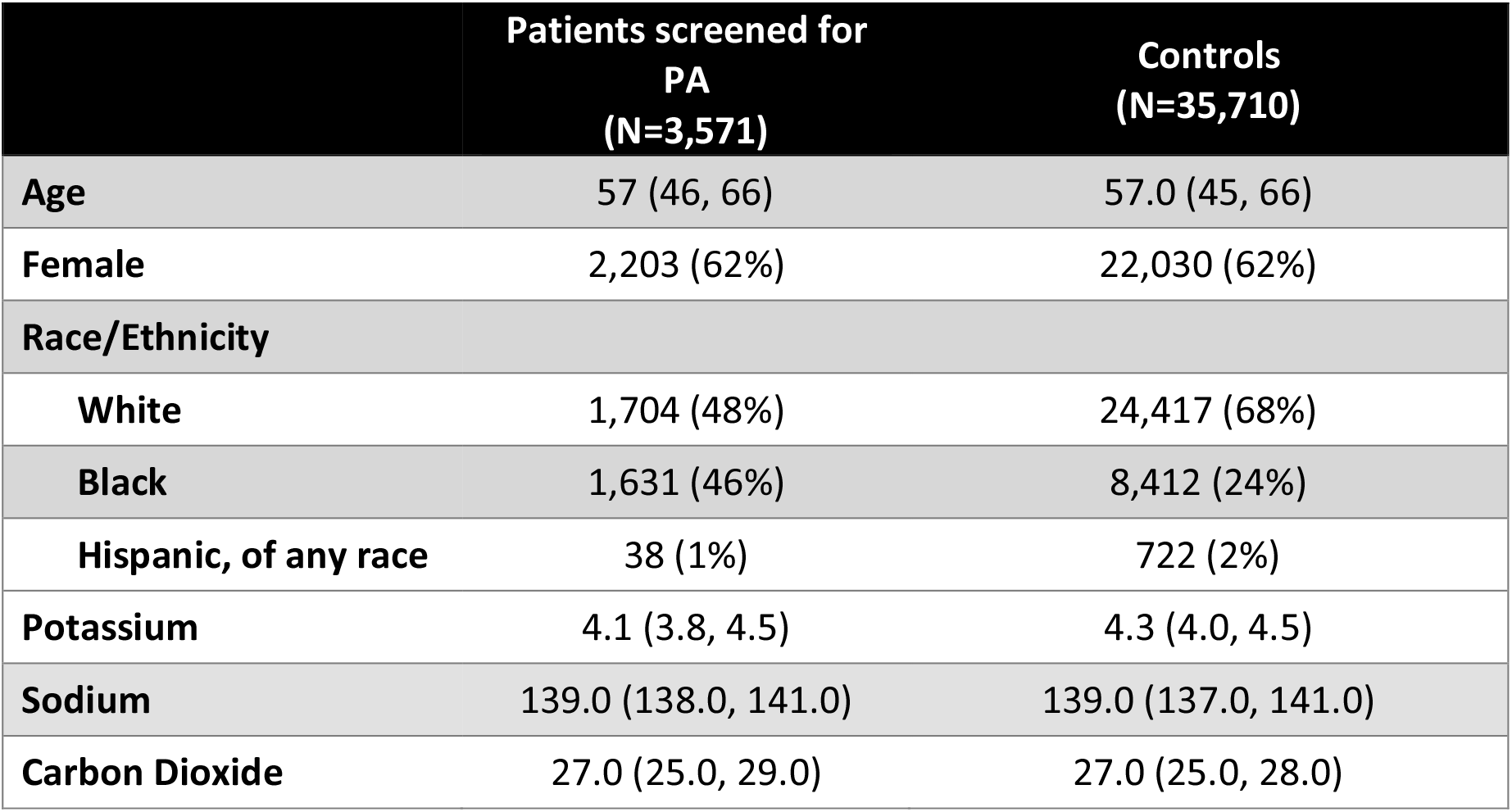
PA Screened and Control Patient Characteristics. EHR demographic and laboratory data for patients with and without paired aldosterone and renin testing, at the time of the evaluation. Numeric results reported as median (quartile 1, quartile 3). Count data reported as # (%).

Based on the expectation that hypokalemia may suppress aldosterone concentrations, we focused on patients with potassium testing at the time of initial PA screening and categorized blood potassium concentrations relative to the test’s reference interval as hypokalemic, normokalemic, or hyperkalemic. At the time of initial PA screening, including a thirty-day window before or after, potassium results were available for 2,940 (82.3%) of patients. Among tested patients, 403 (13.7%) were hypokalemic (≤ 3.5 mEq/L). As expected, this frequency of hypokalemia was considerably higher than that of matched controls (4.3%; OR = 2.9, p < 1.0×10^−10^). In addition, hypokalemic patients were more likely to screen positive for PA (OR = 2.2, p = 1.5×10^−6^).

### Repeat PA Screening

Amongst patients with initial PA screening results that did not meet conservative interpretative criteria, 515 (16%) received repeat screening tests. On first repeat, 7% of those tested positive, which is similar to the positivity rate amongst initial PA screens. Patients with concurrent potassium testing at the time of their initial screen (N=2,682) were overall less likely to undergo repeat PA screening (N=585, OR = 0.5, p = 6.2 ×10^−10^). However, the subset of patients who were found to be hypokalemic (N=340, 13%) were more likely to receive a repeat PA screening testing (23%; OR = 2.1, p = 8.9×10^−7^). Among hypokalemic patients rescreened, 10% (N=8) were positive for PA. Of note, there was a wide range of initial aldosterone concentrations in patients with negative results who were subsequently re-tested (Figure 2).

**Figure 2.**
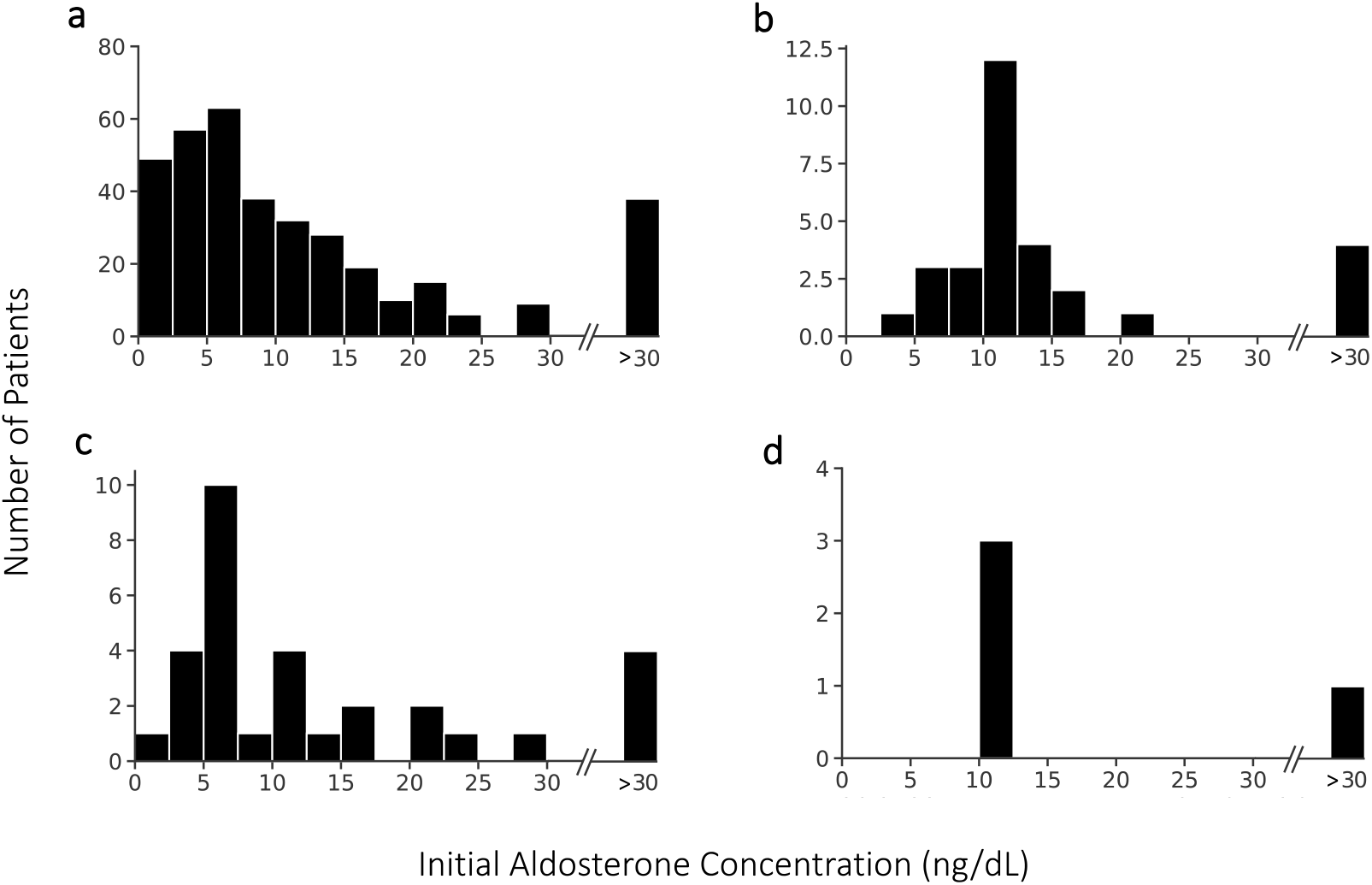
Aldosterone concentrations grouped by repeat PA screening results and corrected hypokalemia. Initial aldosterone concentrations for patients with repeat testing and initial PA screens that did not meet conservative criteria who on subsequent PA screening were (a) negative or (b) positive. Initial aldosterone concentrations for subset of patients with corrected hypokalemia that were (c) negative or (d) positive on repeat.

To investigate the relationship between initial potassium concentration and positivity on repeat, we trained logistic regression models on the 448 initially screen-negative patients with initial potassium testing and repeated PA screening, including 30 patients (7%) who were positive on repeat. Univariate analysis demonstrated an inverse relationship between initial potassium concentration and repeat positivity (OR = 0.57 per SD, p = 0.005).

To better understand the factors associated with positivity on repeat, we sought to describe the patients who underwent repeat screening. Patients re-screened had slightly lower median initial potassium concentrations (4.0 vs 4.2 mEq/L, p = 3.0×10^−4^), higher initial aldosterone concentrations (8.4 vs 7.4 ng/dL, p = 1.9×10^−4^), and lower renin activity (0.8 vs 1.0 ng/mL/hr, p = 0.02). Additionally, they appeared more likely to be identified as Black (48% vs 43%, p=0.05) and were possibly younger (55 vs 57 years, p=0.12). Next, we explored the factors associated with positivity on repeat screening. Patients positive on repeat had a higher median initial aldosterone (11.1 vs 8.0 ng/dL, p = 0.02), lower initial renin (0.3 vs 0.9 ng/mL/hr, p=1.2×10^−5^), and lower initial potassium concentration (3.7 vs 4.1 mEq/L, p = 0.002).

After adjusting for those factors that appear to affect repeat testing or repeat positivity (initial aldosterone concentration, initial renin concentration, age, gender, patient documented race, and the standardized difference in time between repeat tests), the inverse relationship between initial potassium concentration and repeat positivity appeared similar (OR = 0.61 per SD, p = 0.04).

### Hypokalemia and Potassium Repletion

Based on the expectation that potassium repletion among hypokalemic patients increases the sensitivity of subsequent PA screening, we next focused on the patients with repeat PA screening and potassium tests (N=371). The point estimate for the relationship between hypokalemia and repeat positivity (OR = 1.9, p = 0.2) was consistent with the relationship observed between potassium concentrations and repeat positivity. However, we did not see overt evidence of enrichment for PA positivity on repeat testing among patients with corrected hypokalemia (N=35) compared to those who remained hypokalemic (N = 28, OR = 0.77, p = 1.0). Of note, the few patients with corrected hypokalemia who were positive on repeat did appear to have higher initial aldosterone concentrations (Figure 2d).

We next leveraged the longitudinal data to further characterize the covariation of potassium and aldosterone measurements in these patients. We studied 346 patients with repeat PA screening and concurrent potassium tests, excluding patients with hyperkalemia, chronic kidney disease, or changes in medications with large effects on PA screening tests (Figure 1B). Overall, we did not observe a clear relationship between the interval change in potassium concentrations and the change in aldosterone concentrations from initial to subsequent PA screening tests (Slope: 0.98, 95% CI: 0.38 – 1.59; intercept: 3.2 ×10^−5^, 95% CI: - 0.45 – 0.45). However, focusing on patients who were initially hypokalemic and normokalemic on repeat, there was a modest concomitant increase in the median aldosterone concentration (+4.4 ng/dL, IQR: -2.4 – 13.1) compared to patients who were normokalemic for both initial and repeated tests (0.0 ng/dL, IQR: -6.6 – 5.7; p = 6.8×10^−3^; Table 2, Figure 3).

**Table 2.** Changes in Aldosterone and Potassium Concentration According to Change in Potassium Status. Summary statistics for the change in aldosterone for repeated initial ARR tests for potassium trajectory (e.g. hypokalemia to normokalaemia) across repeated tests. The tests of significance are compared to the group of repeated tests that remained normokalemic. Reported as median (Q1-Q3)

|  | Normokalemia to<br>Normokalemia | Hypokalemia to<br>Normokalemia | Hypokalemia to<br>Hypokalemia | Normokalemia to<br>Hypokalemia |
| --- | --- | --- | --- | --- |
| # of paired tests | 223 | 48 | 35 | 40 |
| ΔAldosterone,<br>median (ng/dL) | 0.0<br>(-6.6 – 5.7) | 4.4<br>(-2.4 – 13.1) | 0.0<br>(-6.9 – 3.6) | 0.5<br>(-4.3 – 5.7) |
| *P-value* | - | $6.8 \times 10^{-3}$ | 0.4 | 0.4 |
| ΔPotassium,<br>median (mEq/L) | 0.0<br>(-0.2 – 0.3) | 0.6<br>(0.3 – 0.8) | 0.0<br>(-0.1 – 0.3) | -0.5<br>(-0.8 – (-)0.3) |
| *P-value* | - | $3.7 \times 10^{-16}$ | 0.2 | $7.7 \times 10^{-15}$ |
| ΔTime, median<br>(Days) | 284<br>(49 – 753) | 193<br>(44 – 1092) | 265<br>(24 – 1006) | 435<br>(176 – 1263) |
| *P-value* | - | 0.4 | 0.3 | 0.06 |

**Figure 3.**
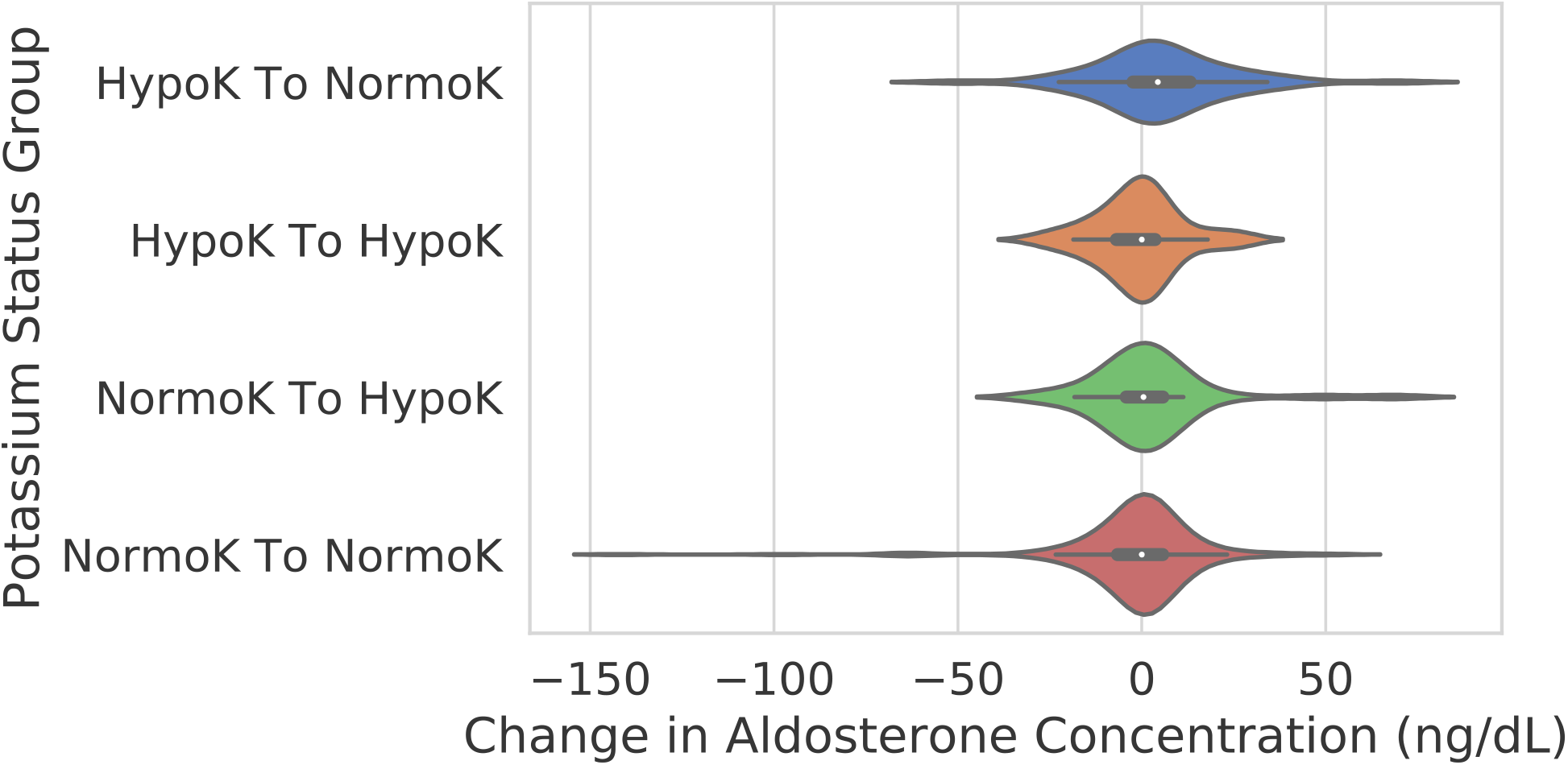
Interval change in aldosterone grouped by change in potassium. Violin plots of the interval change in aldosterone for repeat tests, grouped based on the interval changes in potassium status from initial to subsequent testing. HypoK: Hypokalemia; NormoK: Normokalemia.

To better understand the significance of the observed relationship between the correction of hypokalemia and change in aldosterone concentrations, we further investigated the context of the changes in potassium concentrations. Among all screen-negative hypokalemic patients, only 14 (4.1%) were subsequently prescribed potassium supplementation and continued this up through the time of repeat PA screen and only 4 (1.2%) patients’ potassium appeared corrected to normokalemia. In addition, for these patients there was considerable time between repeat PA screens (Median: 726 days, IQR: 135 – 1,369 days). This interval was similarly long for the subset of patients with corrected hypokalemia (median = 275 days, IQR = 50 – 899 days).

To mitigate the potential effect of interval disease progression in the observed aldosterone elevation, we performed a secondary analysis only including the subset of patients with repeat testing within 2 years (N=245). The median change in aldosterone concentration in the hypokalemic to normokalemic group (+ 4.6 ng/dL) appeared similar and distinct from that of consistently normokalemic patients (0.0 ng/dL, p = 0.04).

## Discussion

PA is underdiagnosed in part due to the complexity of the guidelines for PA screening and their variable and incomplete adoption [27]. Blood potassium concentrations are central to both the pathophysiology of PA and to these screening guidelines. We used in-practice data to assess and reflect upon current PA screening practices in a large, diverse healthcare system, focusing on the measurement of blood potassium as part of PA screening and on the repletion of potassium and repeat PA screening for patients found to be hypokalemic.

By surveying real-world data, we were able to examine the importance of potassium measurement and repletion across the spectrum of PA severity seen in clinical practice. Our patient population appeared similar to that of other published cohorts. We found the frequency of hypokalemia amongst patients being screened for PA was 13%, squarely in between that observed in systematic PA screening cohorts (7 - 9%) and known PA patients (9 - 37%) [5,7,28,29]. In addition, we observed a PA screening positivity frequency of 8.5%, which, as expected, was in between the frequencies observed in unselected hypertension patients (5.9 – 6.1%) and apparent treatment-resistant hypertension (10 – 20%) [5,30].

Current practice guidelines recommend that patients who screen negative for PA and are hypokalemic should undergo repeat screening following potassium repletion. We observed that at the time of initial PA screening, 18% of all patients did not receive concurrent potassium testing, potentially leading to false-negative screening results due to suppression of aldosterone from unappreciated hypokalemia. Additionally, we found that most hypokalemic patients with negative PA screening tests were not rescreened. Even amongst those that were rescreened very few were rescreened following potassium supplementation within the subsequent 2 years. While these patients are more likely to have milder than more severe PA, literature suggests that as a group these patients have excess morbidity [31,32].

Next, to complement the classic *in vivo* experiments that form the basis for the current recommendation, we examined the apparent impact of blood potassium on PA screening results in practice [17,18,23]. Amongst patients with repeat PA screens following an initial negative screen, we found that a one standard deviation lower initial potassium concentration (3.5 versus 4.1 mEq/L) was associated with a 75% higher odds of testing positive for PA on repeat testing. This represented an increase in absolute repeat PA positivity frequency from 5.9% to 9.9%. While there was selection bias affecting who underwent repeat PA screening, the increased odds of repeat screening positive for PA did not appear to be explained in a large part by initial aldosterone and renin results, demographics, and time between tests. This finding supports the guidelines’ recommendation for universal potassium testing and repeat PA screening if hypokalemic.

If we were to implement universal potassium testing and repeat PA screening in our PA screening practice, we would expect approximately 1 in 8 screened patients to be hypokalemic and approximately 10% of hypokalemic patients to be positive on repeat. Thus, among initially negative patients (92%) we would expect approximately 1 in 80 (1.1%) of all screened patients to be hypokalemic and PA positive on repeat. If applied initially to all patients in our population, this would correspond to an 11% relative increase in patients identified with likely PA and an absolutely increase in the initial positivity rate from 8.5% to 9.6%.

We examined how such a universal a potassium testing and rescreening program compares to our current practice. We found that 0.2% of all patients in our population were initially hypokalemic and positive for PA on first repeat. Compared to a universal potassium screening protocol, our current screening practice is identifying far fewer positive patients on repeat. Based off our prior estimates, our current screening practice is missing about 80% of potential patients with likely PA on repeat. While this gap could be explained by our estimates being biased by unmeasured determinants of screening that affect positivity on repeat, given the low numbers of hypokalemic patients with repeat testing or potassium supplementation, and the wide range of initial aldosterone concentrations we do not believe that our population was disproportionately enriched.

The potential benefits of increasing the frequency of diagnosis of primary aldosteronism must be balanced with the added costs of additional screening. Without targeted therapy or adrenalectomy, these patients on average have considerably higher mortality and morbidity [1–6]. While patients with relatively low aldosterone concentrations are unlikely to currently have severe PA, literature suggests that even milder forms of PA confer excess morbidity and can develop into severe forms of the disease [31,32]. Additional studies are needed to understand the benefit of targeted treatment in this specific subset of mild PA patients [33–35].

Rescreening all hypokalemic patients for PA is reasonable as the positivity rate we observe in repeated hypokalemic patients (~10%) is consistent with that of other patient groups recommended by guidelines to be screened. However, retesting could be further limited to patients with somewhat elevated aldosterone concentrations to moderate the cost of implementation and improve repeat positivity [7]. We examined the potential impact of restricting repeat screening based on aldosterone concentrations. Only retesting patients with initial aldosterone concentrations > 5 ng/dL (N=53) corresponded to an apparent repeat positivity frequency of 15% and included 100% of patients who had been subsequently screen-positive. Further restricting rescreening to aldosterone concentrations >10 ng/dL (N=36), the repeat positivity frequency would have been 31% and we would have identified only 83% of patients who were subsequently positive. The observed improved repeat positivity with aldosterone thresholds must be balanced against the potential loss of sensitivity and the effort needed to operationalize this in practice.

Based on these findings, we recommend building operational processes and tools to support PA screening workflows in order to increase PA screening yield and thereby mitigate and decrease the associated morbidity and long-term healthcare costs [35,36]. One potential route to enable more consistent identification of hypokalemic patients would be using a PA screening order set that includes measurement of blood potassium concentration. Ideally, these screening tests’ results would include a rich interpretive comment to support screening interpretation, in particular noting that non-diagnostic results should be considered indeterminate for patients with hypokalemia and high aldosterone [37].

While this work reveals large numbers of patients potentially affected by known and unknown hypokalemia, we recognize several limitations. This is a retrospective study of real-world data and we were not able to identify and account for all potential sources of confounding. In particular, we were not able to fully adjust for why patients were screened, why some patients were rescreened, or why there were interval changes in potassium concentrations. Additionally, we have limited data on the relationship between changes in aldosterone and potassium, because few hypokalemic patients appeared to have been corrected to normokalemia expressly for the purpose of repeat PA screening. Thus, we were unable to explicitly demonstrate that potassium repletion improves repeat positivity. This highlights the need for future prospective studies further examining the relationship between aldosterone and potassium correction.

## Data Availability

Restrictions apply to the availability of some or all data generated or analyzed during this study to preserve patient confidentiality. The corresponding author will on request detail the restrictions and any conditions under which access to some data may be provided.

## Acknowledgments

The authors would like to thank Imran Ajmal, Paul Lee, and Thaibinh Luong for their technical expertise and support.

## Data Availability

Restrictions apply to the availability of some or all data generated or analyzed during this study to preserve patient confidentiality or because they were used under license. The corresponding author will on request detail the restrictions and any conditions under which access to some data may be provided.

